# Head-to-head containment performance testing of four commercially available closed-system transfer devices using the 2016 Draft NIOSH test protocol and 2.5% w/v 2-phenoxyethanol as challenge agent

**DOI:** 10.64898/2026.07.21.26358603

**Authors:** Alan-Shaun Wilkinson, Kate Walker, Laima Ozolina, Romana Bon, Andrew Wallace, Michael Charles Allwood

## Abstract

**Objectives:** The 2016 Draft National Institute for Occupational Safety and Health (NIOSH) universal protocol “A Performance Test Protocol for Closed System Transfer Devices Used During Pharmacy Compounding and Administration of Hazardous Drugs.” employing ATD-GC-MS with the 2016 Draft NIOSH proposed challenge agent 2-Phenoxyethanol (2-POE, 2.5% w/v in water) was used to assess the containment performance of two barrier and two air-cleaning CSTDs in a head-to-head test with replication (n=4) for both NIOSH tasks 1 (drug preparation) and 2 (drug administration). The study objective was to assess the containment performance of both physical barrier and air-cleaning CSTDs.

**Methods:** ChemoLock™ barrier (ICU Medical, USA), ChemoLock™ air-cleaning (ICU Medical, USA), PhaSeal™ barrier (Becton Dickinson, USA), and Tevadaptor™ air-cleaning (Simplivia, IL – now marketed as Chemfort™) CSTDs were evaluated using the 2016 Draft NIOSH universal test protocol for Task 1 (n=4) and Task2 (n=4). A 100 mL empty IV bag, accessed with a CH-10 ChemoClave™ bag spike was used with the IV administration set in Task 2 to avoid pressurisation of the occluded administration set during the simulated “IV push”. Air samples were collected (GilAir pump) at 100mL/minute for 30 minutes on Tenax TA thermal desorption (TD) tubes (Markes, UK). 2-POE, a semi-volatile surrogate mimicking hazardous drugs was prepared at 2.5% w/v solution in MilliQ water. This was aliquoted (50mL) into drug vials and crimp sealed with a butyl rubber stopper (Adelphi, UK). 2-POE was analysed against a calibration curve (n=8) in the range 0.6ng to 200ng using analytical methods published previously. Chromeleon v7.3 CDS (Thermo Scientific, UK) was used for analysis based on detection of SIM ions (M/Z: 77, 94, 138). Blank TD tubes (n=85) were used to determine the experimental LOD and LLOQ. Negative control tests (employing MilliQ water as surrogate) were performed for each CSTD Task 1 (n=1) and Task 2 (n=1). An open “needle and syringe” positive control test was performed at 5 and 10 microlitre volumes in the NIOSH chamber to demonstrate system suitability.

**Results:** Chamber cleaning and analysis performed between tests, gave average blank values (n=85) of 0.05±0.01 ppbv (95% confidence interval). The experimental limit of detection (LOD) and quantification (LLOQ) for the tests was 0.16±0.01 ppbv and 0.41±0.01 ppbv respectively. The positive control tests produced signals of 3.13 ppbv (n=1) and 6.59 ppbv (n=1) for 5 microlitre and 10 microlitre releases of 2-POE respectively and demonstrated system suitability to quantify down to a 650 nanolitre liquid release volume. (1) ChemoLock™ barrier, (2) ChemoLock™ air-cleaning, (3) BD PhaSeal™ barrier and (4) Tevadaptor™ air-cleaning CSTDs all generated <LLOQ signals for both NIOSH tasks 1 and 2. No statistically significant difference was found between the containment performance of the two air-cleaning and the two physical barrier CSTDs evaluated in this study.

**Conclusions:** System suitability was demonstrated using deliberate releases of the 2-POE surrogate at 5 and 10 microlitre volumes producing 2-POE concentrations in the NIOSH chamber of 3.13 ppbv and 6.59 ppbv respectively. Evaluation of the containment performance gave <LLOQ (0.41±0.01ppbv) for all four of the CSTDs tested in the study: (1) ChemoLock™ barrier, (2) ChemoLock™ air-cleaning, (3) BD PhaSeal™ barrier and (4) Tevadaptor™ air-cleaning, based on testing of four replicates of each technology in both NIOSH Task 1 and NIOSH Task 2. There was no statistically significant difference in containment found between the two air-cleaning and the two barrier CSTDs evaluated in this study.

## Introduction

### Background

Many anti-cancer drugs provide potentially therapeutic benefits to patients whilst posing a hazard to healthcare workers through unintended occupational exposure [1,2]. Healthcare workers exposed to anti-cancer drugs may exhibit a range of acute symptoms including headache, allergic reactions, and nausea and vomiting, or long-term effects such as genotoxicity, infertility, and fetal abnormalities [2]. These risks prompted the publication of guidelines in the United States (US) by the NIOSH and internationally by the Health and Safety Executive (HSE) in the UK for the safe handling of cytotoxic and other hazardous drugs [2–6]. Use of primary engineering controls such as a ventilated cabinet or isolator and personal protective equipment (PPE) are required for the preparation of hazardous drugs, but the use of a closed system transfer device (CSTD) [1] is currently only recommended as an additional hierarchical layer of operator protection during drug preparation [1,7]. CSTDs utilise a wide variety of containment technologies and it is therefore necessary to assess the containment of each CSTD technology using a scientifically robust methodology. With the recent publication of USP <800> there is a requirement to use CSTDs for hazardous drug administration when the dosage form allows [7]. USP <800> recognises the need to evaluate the performance of CSTDs *via* peer-reviewed studies as well as reduction in contamination [7]. When compounding hazardous sterile drugs, both the recently updated USP <797> and USP <800> standards apply, which are the respective controls for sterility and containment [7,8]. USP <800> requires the use of CSTDs for the administration of all hazardous drugs and recommends their use for the preparation of hazardous drugs that are on the hazardous drug list published by the NIOSH [8,9]. Implementation of a CSTD provides an additional layer of protection on the ward where engineering controls are not present, reducing reliance upon PPE alone and offering improved protection for healthcare workers on the ward [2,3,10]. The NIOSH recommends using CSTDs to limit occupational exposure to hazardous drug materials and sharps when compounding and administering hazardous drugs [1].

The NIOSH defines a CSTD as a drug transfer device that “mechanically prevents the transfer of environmental contaminants into the system and the escape of hazardous drug or vapor concentrations outside the system” [1]. CSTDs may reduce the risk of needle-stick injuries, spillages of hazardous drug materials and the formation of aerosols during the compounding and administration processes, which are common sources of contamination [3,10]. CSTDs can help to reduce exposure by containing liquid, aerosol and vapour releases [1]. This is achieved by employing either a physical barrier that captures the displaced air which originated from within the hazardous drug vial, or by the use of an air-cleaning technology that specifically cleans the air that passes between the hazardous drug vial and the environment during the pressure equalisation step [1,11]. Using either system, helps to protect the operator from accidental exposure to the hazardous drug material and also helps protect the drug product from external microbiological contamination.

In 2015 the NIOSH published a draft protocol to assess the vapour containment performance of physical barrier CSTDs in response to requests from within the healthcare industry for an independently developed containment performance test protocol [12]. The 2015 Draft NIOSH test protocol employed 70% isopropyl alcohol (IPA) as the test surrogate [12]. In January 2016 the NIOSH published a request for information to assist in the development of a new draft CSTD protocol to test the containment performance of air-cleaning CSTDs [13]. Wilkinson *et al.* in collaboration with the UK Health and Safety Executive (HSE), submitted information to the federal docket in response to the request for information (RFI) [14]. Wilkinson *et al.* provided a methodology that enabled testing of both physical barrier and air-cleaning CSTDs [14]. This provided a blueprint for a universal test protocol for the assessment of containment performance of CSTDs regardless of the technology that they employ [14]. In September 2016, the NIOSH released the 2016 Draft NIOSH universal test protocol for the assessment of the containment performance of both physical barrier and air-cleaning CSTD technologies [15] which was based on the protocol information submitted by Wilkinson *et al.* [14]. The 2016 Draft NIOSH protocol allowed for the first time a quantitative assessment of the containment performance of all types of CSTD irrespective of the technology employed, either physical barrier, air-cleaning or other (future technologies not yet available on the market) [15]. The 2016 Draft NIOSH protocol differs from the 2015 Draft NIOSH protocol in that it does not generate a real time measurement but instead produces a time weighted average (TWA) collection of the surrogate on to a sorbent tube which is then analysed immediately after completion of the test. Collection of the surrogate released is performed from within a closed chamber avoiding the possibility of dilution of any released surrogate material in the NIOSH chamber [14–15]. The 2016 Draft NIOSH test protocol is aligned with the UK Health and Safety Executive (HSE) method for the assessment of volatile organic compounds using sorbent tubes (mdhs104), solvent desorption or thermal desorption and gas chromatography [16] as well as method developed by the US Occupational Safety and Health Administration (OSHA) for monitoring levels of volatile and semi-volatile organic materials in the environment [16–19]. The Draft 2016 NIOSH protocol states that all surrogates used in the testing of CSTDs should have a vapour pressure of at least that of thiotepa (1.0 × 10^−2^ mm Hg) and up to 1 mm Hg and thus provide a safety factor of at least one hundred over the most volatile hazardous drug (based on thiotepa) [15].

The reader can find more information on the background to the development of the 2016 Draft NIOSH universal test protocol for CSTDs along with a Q and A section on the CDC NIOSH website for Closed System Drug-Transfer Device (CSTD) research page, this also discusses ongoing research being conducted at the CDC NIOSH [20].

Since USP <800> [7] there has been a paucity of peer reviewed scientific studies published using the latest 2016 Draft NIOSH universal CSTD test protocol with Wilkinson *et al.* being the only authors to date to publish experimental containment data obtained using the 2016 NIOSH Draft test protocol as applied to the containment performance of CSTDs employing different technologies [21–22]. Given the requirement of USP<800> that CSTDs must be used for administration of hazardous drugs there is an increasing need for the generation of containment performance data for CSTDs to be published in independent peer reviewed scientific studies that employ the latest 2016 Draft NIOSH universal test protocol [15]. At the time of writing there are only two published studies that employ the 2016 Draft NIOSH methodology and NIOSH surrogates as applied to different types of CSTDs [21,22]. Empirical containment data is critically important for the assessment of both physical barrier and air-cleaning CSTDs to inform selection and purchasing decisions for which CSTD to use on the ward [7,15]. Furthermore, because lower cost CSTDs may provide a route to health worker protection in a variety of situations, there is a defined benefit to healthcare organisations to acquire and compare containment performance data covering all current CSTD technology platforms [7].

The present study aims to provide quantitative containment data for two physical barrier and two air-cleaning CSTDs in the same test cohort thereby allowing direct comparisons in containment performance to be made between the two disparate CSTD technologies [15]. This study exploits the potential of the 2016 Draft NIOSH universal performance test protocol in the assessment of CSTD containment performance across different technologies using the published NIOSH surrogate of 2.5% w/v 2-phenoxyethanol (2-POE) in water adding further scientific data to that previously published [15,21–22]. Wilkinson *et al.* previously used the same NIOSH challenge agent 2-POE but only evaluated the containment performance of a single air-cleaning CSTD device type [21,22]. Subtle changes to the design and manufacture of the NIOSH environmental chamber and setup were previously published by Wilkinson *et al.* to support more robust quantitation of releases of the NIOSH challenge agent [23]. In the present head-to-head study, we report the containment performance of two physical barrier: (1) ChemoLock™ (ICU) and (2) Phaseal™ (BD); and two air-cleaning CSTDs: (3) ChemoLock™ (ICU) and (4) Tevadaptor™ (Simplivia) using the NIOSH challenge agent 2.5% w/v 2-phenoxyethanol (2-POE) in water.

All manipulations of CSTDs were performed according to the manufacturer’s instructions for use (IFU) during execution of the 2016 Draft NIOSH universal CSTD test protocol to prevent generation of invalid results.

In the 2016 Draft NIOSH universal test protocol nine potential surrogates were suggested including 2-POE based on their vapour pressure values and the reader is referred to the 2016 Draft NIOSH protocol document for more detailed information on surrogate selection [15]. Whilst experimental data on vapour pressure is available for a limited selection of hazardous drugs [24], when no data exists there is a requirement to estimate vapour pressure values using in-silico methods such as those provided by the EPI suite of programs [25]. The EPI software was developed by the US Environmental Protection Agency (EPA) [25]. The present study employed the NIOSH surrogate 2-POE which is one of nine surrogates proposed by NIOSH based on their vapour pressure values relative to the vapour pressure of the most volatile hazardous drug thiotepa with an additional safety factor of one hundred [15]. The selection of 2-POE as a surrogate material is also cited as an appropriate semi-volatile surrogate and employed in published scientific works by Pengelly *et al.* of the UK Health and Safety Executive (HSE) [26]. Pengelly *et al.* highlights concerns over the uncertainty and potential for errors when estimating exposure risk of humans to the vapour from hazardous chemical materials when this is based on published saturated vapour concentration (SVC) values [26]. Pengelly *et al*. reports the relationship between the fundamental physiochemical property of SVC and measured airborne concentrations of surrogates in low volatility chemical substances by use of appropriate candidate compounds such as 2-POE [26]. Pengelly *et al.* compared both small-scale baseline and larger volume scale experiments using the same NIOSH surrogate 2-POE designed to accurately mimic real world worker exposure environments using a specially designed test room [26]. Importantly Pengelly *et al.* demonstrated that semi-volatile materials with low saturated vapour pressure (SVP) of typically 0.7 × 10^−2^ mm Hg and lower generate significantly less chemical vapour in real world tests than would be predicted by calculation using the saturated vapour pressure or SVC values and generally only about 1% of their theoretical saturated vapour concentration (SVC) values [26]. Therefore, according to Pengelly *et al.* use of the theoretical SVC data in predicting airborne concentrations of low volatility hazardous materials will result in significantly overestimation of the exposure risk as was found using the NIOSH surrogate 2-POE in real-world large scale studies [26].

According to Pengelly *et al.* volatile surrogates, for example isopropyl alcohol (IPA) used in the 2015 Draft NIOSH test protocol are likely to achieve significant chemical vapour concentrations approaching 100% of their SVC values when released or placed in an open container [26]. The high concentration of IPA that can be achieved in the vapour when used as surrogate is therefore significantly different to what has been observed and reported by Pengelly *et al.* when equivalent releases are performed using the semi-volatile chemical compound 2-POE as the NIOSH surrogate [26]. This result generates around another factor of one hundred increase in concentration when IPA is used as surrogate in addition to the safety factor of the surrogate having one hundred times higher vapour pressure than the most volatile hazardous drug, as required in the 2016 Draft NIOSH universal test protocol [15]. Wheeler *et al.* performed real world studies to assess the release of vapour from two hazardous materials, the pesticides prallethrin and bioallethrin [27]. Both of these hazardous substances failed to generate airborne concentrations that were greater than 0.6% of their stated SVC even when directly heated to increase evaporation [27]. Other data obtained using both ATD-GC-MS and selected-ion flow-tube mass spectrometry (SIFT-MS) by the NIOSH, HSE and Element (a UK distributor or SIFT-MS technology formerly Anatune) measured the airborne concentrations of the NIOSH surrogate 2-POE from liquid releases in a closed chamber (including NIOSH chamber) and reported these to be typically a few percent of the predicted concentration in the vapour based on the predicted SVC value for 2-POE [28,31]. Reported vapour concentrations for the NIOSH surrogate 2-POE are typically a few precent of the predicted value based on SVC and this appears to be consistent for other semi-volatile materials and potentially may also be the case for real hazardous drug materials.

The 2015 Draft NIOSH test protocol employing IPA as a surrogate remains a popular choice as can be seen by the number of recent publications that employed this methodology [32–37]. The present study will add to the body of scientific containment performance data from adoption of the latest 2016 Draft NIOSH universal test protocol in combination with the NIOSH surrogate 2-POE [15]. It should be noted that there are nine NIOSH surrogates proposed in the 2016 Draft NIOSH universal test protocol of which 2-POE is one [15]. The selection of 2-POE was made for the present study based on the body of work performed by Pengelly *et al.* at the UK Health and Safety Executive (HSE) on this NIOSH surrogate [26]. To our knowledge there is no comparable real world test data available for the other eight NIOSH surrogates suggested in the 2016 Draft NIOSH universal test protocol for liquid release and evaporation to produce airborne chemical concentrations in the vapour [15].

In summary, the rationale for the current study was to employ the 2016 Draft NIOSH proposed challenge agent 2-POE and use the latest 2016 Draft NIOSH protocol methodology to evaluate the containment performance of four commercially available CSTDs including two physical barrier and two air-cleaning CSTDs. The null hypothesis for the study is that the CSTDs under test are not able to completely contain the 2-POE surrogate whilst carrying out NIOSH Task 1 and Task 2. The signal from release of 2-POE for the CSTD under test provides a direct measure of containment (or lack of) of the CSTD. The higher the signal of 2-POE in parts per billion volume the poorer the CSTD is at containing the challenge agent 2-POE which is a marker for the containment of real hazardous drugs.

## Materials

2.5% w/v 2-POE was prepared by addition of 2.5 grams of 2-POE (Sigma Aldrich, 90% or better, CAS No. 122-99-6) and made up to 100 mL in a volumetric flask (Grade A) (Fisher, UK) using Milli-Q water (Integral system, Merck Millipore, UK), mixed by shaking, and 50 mL aliquoted into 100 mL type I glass vials (Adelphi, UK). Negative control 100 mL drug vials (Adelphi, UK) were prepared by directly adding 50 mL of Milli-Q water, and then sealed. Stainless steel inert coated thermal desorption (TD) tubes 89 mm long × 6.4 mm outer diameter and packed with 200 mg of Tenax TA (Markes, UK) were used throughout the study. Internal standard was achieved by accurately weighing out d8 toluene (Sigma Aldrich, UK) and dispersing in five nines analytical grade di-nitrogen (BOC, UK) to prepare a 1ppm standard. The internal standard was then added to each tube immediately prior to analysis using the Markes instrument control (MIC) software. Calibration of the instrument was performed by direct spiking of microlitre liquid aliquots of 2-POE standards prepared in LCMS grade methanol (Fisher, UK) on the Tenax tubes (Markes, UK) over the range of 0.6ng to 200ng mass loading per tube. Tubes containing calibration (CAL) standards at eight CAL levels were analysed using internal standards in an identical manner to the test and blank tubes and a full calibration curve was performed within each sequence of test tubes. The details for all closed system components used in the testing are detailed in table 1 below.

**Table 1.** Table showing CSTD device components tested in the study during Task 1 and Task 2.

| Number | Task | CSTD Type | Component(s)<br>Part Number | Lot Number | Expiry |
| --- | --- | --- | --- | --- | --- |
| 1 | Task 1 | ChemoLock™<br>Barrier<br>(ICU Medical) | CL2000S | 5438215 | 01-Jun-2026 |
|  |  |  | CL-80S | 5236951 | 01-Mar-2026 |
|  |  |  | CL-12 | 5329899 | 01-Apr-2026 |
| 2 | Task 1 | ChemoLock™<br>Air-Cleaning<br>(ICU Medical) | CL2000S | 5438215 | 01-Jun-2026 |
|  |  |  | CL-70S | 5166676 | 01-Feb-2026 |
|  |  |  | CL-12 | 5329899 | 01-Apr-2026 |
| 3 | Task 1 | Tevadaptor™<br>Air-Cleaning<br>(Simplivia™, IL) | 412111 | UA1536 | 31-Aug-2023 |
|  |  |  | 412126 | UAA521 | 31-Sep-2023 |
|  |  |  | 412113 | UA3528 | 30-Sep-2023 |
| 4 | Task 1 | BD PhaSeal™<br>Barrier<br>(Beckton<br>Dickinson) | 515105 | 1911114 | 31-Oct-2024 |
|  |  |  | 515306 | 2005214 | 30-Apr-2024 |
|  |  |  | 515003 | 1910105 | 31-Mar-2022 |
| 5 | Task 2 | ChemoLock™<br>Barrier<br>(ICU Medical) | CL2000S | 5438215 | 01-Jun-2026 |
|  |  |  | CL-80S | 5236951 | 01-Mar-2026 |
|  |  |  | CL-12 | 5329899 | 01-Apr-2026 |
|  |  |  | CL2100 | 5246838 | 01-Mar-2026 |
| 6 | Task 2 | ChemoLock™<br>Air-Cleaning<br>(ICU Medical) | CL2000S | 5438215 | 01-Jun-2026 |
|  |  |  | CL-70S | 5166676 | 01-Feb-2026 |
|  |  |  | CL-12 | 5329899 | 01-Apr-2026 |
|  |  |  | CL2100 | 5246838 | 01-Mar-2026 |
| 7 | Task 2 | Tevadaptor™<br>Air-Cleaning<br>(Simplivia, IL) | 412111 | UA1536 | 31-Aug-2023 |
|  |  |  | 412126 | UAA521 | 31-Sep-2023 |
|  |  |  | 412113 | UA3528 | 30-Sep-2023 |
|  |  |  | 412114 | UA4519 | 31-Aug-2023 |
| 8 | Task 2 | BD PhaSeal™<br>Barrier<br>(Beckton<br>Dickinson) | 515105 | 1911114 | 31-Oct-2024 |
|  |  |  | 515306 | 2005214 | 30-Apr-2024 |
|  |  |  | 515003 | 1910105 | 31-Mar-2022 |
|  |  |  | 515202 | 1910109 | 30-Sep-2023 |

### Equipment

A NIOSH chamber was constructed as described previously by Wilkinson *et al.* [21,23]. The chamber was sealed during testing using hand-spring clamps applied to the flexible tubing (Tool station, UK). Three calibrated air sampling pumps (Gil-Air, UK) were used, two to sample the chamber air for both blank and test measurements and one to collect the environmental background air on each day of test. TD tubes were conditioned using the same Markes TD100-xr instrument as used for analysis of tubes prior to use. An analytical standard of d8 toluene (Sigma Aldrich, UK) was used for preparation of internal standard for addition to TD tubes. This was achieved by accurately weighing out d8-toluene liquid with dissolution in research grade di-nitrogen gas (BOC, UK) to provide a 1ppm standard. An external B105 diaphragm pump (Charles Austin, UK) was used to purge the chamber with clean air prior to use. Sample analysis was performed using an automated Thermo Scientific Trace 1300 gas chromatograph fitted with a TD100-xr thermal desorption autosampler (Markes, UK) interfaced to a Thermo Scientific ISQ 7000 AEI mass spectrometer. The instrument performance was checked each day using an automated tune check followed by a tune of the mass spectrometer if required. Separation was achieved using a VOCOL capillary column (60 m length × 0.25 mm diameter × 1 µm film thickness) (Sigma-Aldrich) and instrument control, data acquisition and analysis was performed using the CDS software supplied by Thermo Scientific (Chromeleon version 7.3).

## Methods

Full methodology details have been published previously, please refer to the supplemental materials provided electronically [S1 Text][21].

The assessment of CSTD containment performance was performed in accordance with the 2016 Draft NIOSH universal CSTD test protocol [15] as follows:

- A 2.5% w/v POE solution in Milli-Q water was used as the challenge agent
- An air purge of the chamber was performed at a flow rate of 15 L/min for 30 minutes prior to use.
- An environmental air sample was taken each day prior to testing.
- Air samples were collected at a flow rate of 100 mL/min and for 30 minutes using dual air sampling pump systems connected to stainless steel inert coated Tenax TA TD tubes.
- Dual blank chamber measurements were recorded prior to testing of a CSTD to ensure no contamination inside the chamber.
- A validated chamber cleaning procedure was employed throughout the testing to ensure low background levels for 2-POE.
- All testing procedures were performed strictly in accordance with manufacturer instructions for use (IFU) for each CSTD technology evaluated to ensure validity.

For each measurement (blank and test) dual air sampling pumps were used to collect duplicate air samples (n=2). NIOSH tasks 1 and 2 were performed in replicate (n=4) for both Task 1 and Task 2 for each CSTD technology assessed as stated in the 2016 Draft NIOSH universal CSTD test protocol except for the improvement made to Task 2 by addition of a closed “empty” 100mL IV bag to collect the IV push [15]. The addition of an IV bag waste receptor did not impact on the results and if anything reduced the likelihood of over-pressurisation of the IV bag containing the diluent when making the IV “push” compared with the original setup for performing NIOSH Task 2. Negative controls were performed using 100% Milli-Q water as challenge agent for each of the NIOSH tasks 1 (n=1) and 2 (n=1) for each CSTD evaluated to demonstrate system suitability. Positive controls were performed using a needle and syringe “open system” for release of liquid aliquots of the 2.5% w/v 2-POE in water challenge agent at two volume release levels: 5 and 10 microlitres to demonstrate system response and suitability. The selection of a surrogate 2-POE concentration of 2.5% w/v has been discussed by the authors previously [21] and represents a clinically relevant concentration of hazardous drug whilst maintaining adequate solubility for the surrogate in aqueous solution. It is also a concentration of surrogate that has been demonstrated to have no chemical incompatibility issues with different plastics employed in various medical devices including CSTDs. In contrast the authors have observed significant chemical incompatibility issues with a range of polymeric materials and medical devices when used in conjunction with the surrogate 70% IPA (unpublished data). Data is however freely available in the open literature on chemical compatibility between various polymeric materials used in medical devices and a range of organic solvent materials [38].

### Sample analysis

Sorbent tubes were analyzed using automated ATD-GC-MS detection with the MS operating in selective ion mode (SIM) using two qualifier ions and one ion for quantitation for both the internal standard d8-toluene and 2-POE, details of which have been published previously [21]. Analytical conditions and instrument calibration are described fully in the supplemental materials [S1 text]. A time weighted average (TWA) sample approach allowed for determination of the mass of 2-POE collected on the sorbent tubes, in nanograms, against a standard calibration curve. An example of a typical calibration curve for 2-POE is shown in figure 1 below. The mass of 2-POE was then converted to an airborne concentration in the test chamber in ppbv following the procedures stated in HSE MHDS104 [16]. The calibration data was fitted using a factor 2 polynomial equation with correlation R^2^ of at least 0.999 in accordance with ICH guidelines requirements (R^2^ of 0.99) and methods were compliant with the ISO 16017-1:2000 requirements for indoor, ambient and workplace air sampling and analysis [39–41].

**Figure 1.**
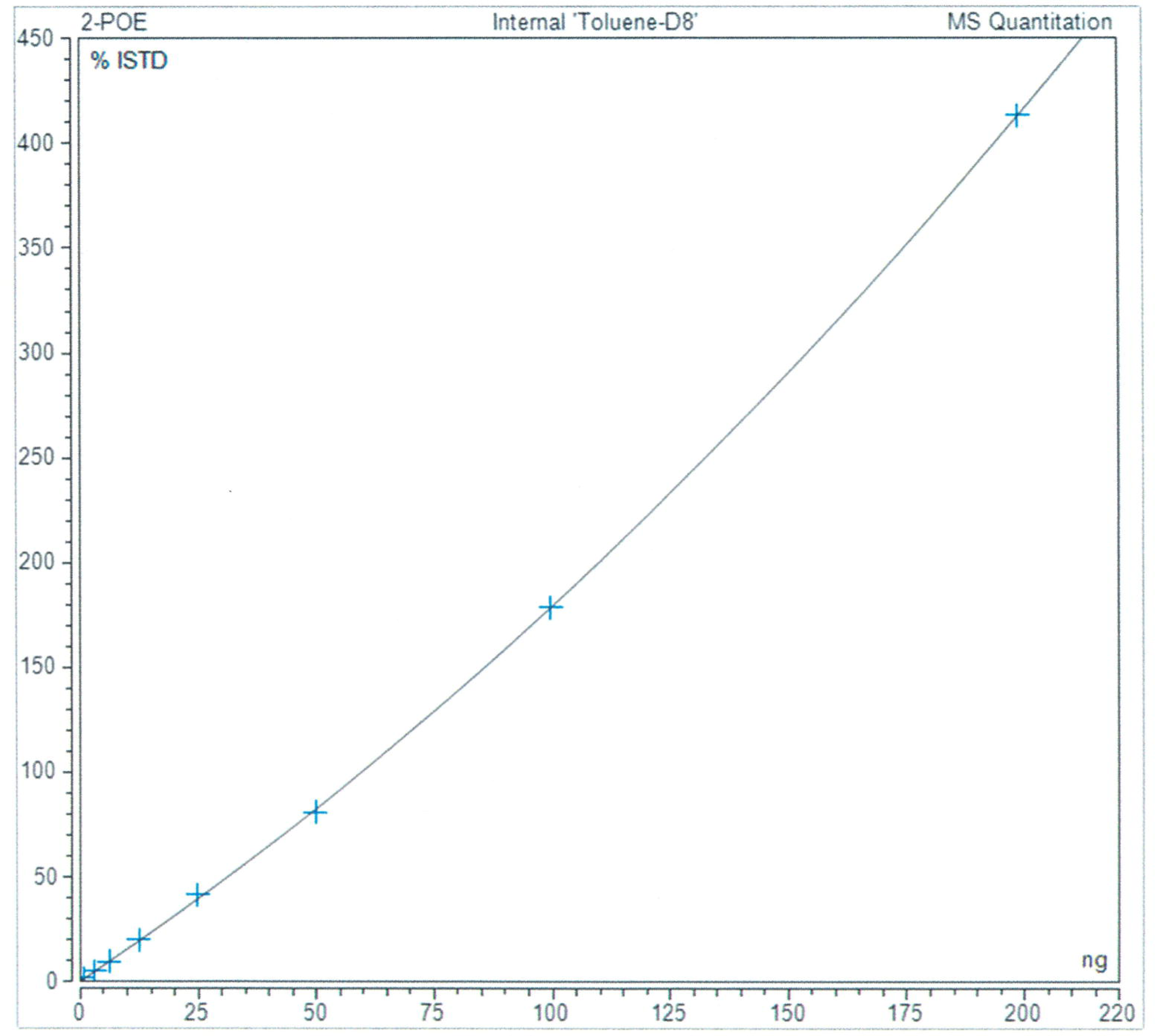
Calibration plot for 2-phenoxyethanol by automated thermal desorption gas chromatography mass spectrometry (ATD-GC-MS).

In addition to the analytical instrumental calibration shown above in figure 1 it was important to calibrate for liquid releases of 2-POE from the challenge agent solution using known volumes of release of the 2.5% w/v 2-POE surrogate solution in the chamber. This was performed at 5 microlitre and 10 microlitre volume release values. A third known value was also included for the mean environmental background concentration for 2-POE from chamber blank measurements. The results are plotted in figure 2 below showing the relationship between volume of challenge agent solution released versus signal for 2-POE in ppbv.

**Figure 2.**
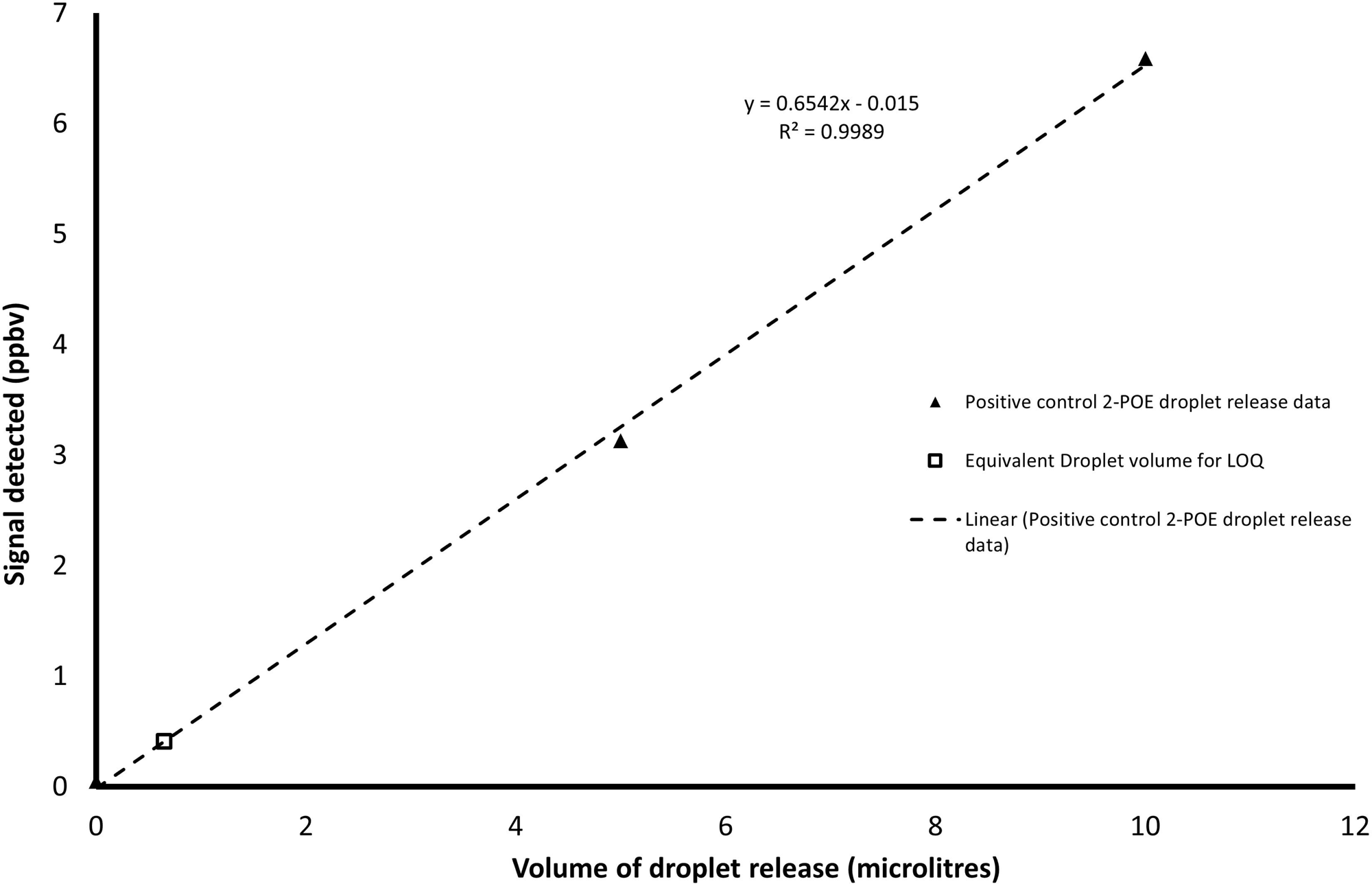
Calibration plot for liquid releases of 2.5% w/v 2-phenoxyethanol in the NIOSH chamber using known volume microlitre releases of 2.5% w/v 2-phenoxyethanol.

The correlation between liquid volume release and signal was 0.999 showing excellent correlation between these two parameters [40,41]. The open square marker in figure 2 above shows the smallest liquid volume of release that can be quantified using the validated analytical method and is based on the LLOQ of 0.41 ppbv at 650 nanolitres of challenge agent solution.

Data processing, determination of the experimental LOD and LLOQ, and data analysis were performed in accordance with the HSE MHDS104 method [16]. The POE release values are reported without background subtraction of the blank samples recorded immediately prior to the test. No carryover of 2-POE was observed in the chamber following CSTD testing, hence no additional cleaning of the chamber was required between tests. The implementation of a robust cleaning procedure ensured low background values for 2-POE and a commensurate low LLOQ was maintained throughout the testing. Figure 3 shows the BSTL modified NIOSH chamber. All changes to the original NIOSH chamber are documented and described elsewhere [21,23].

**Figure 3.**
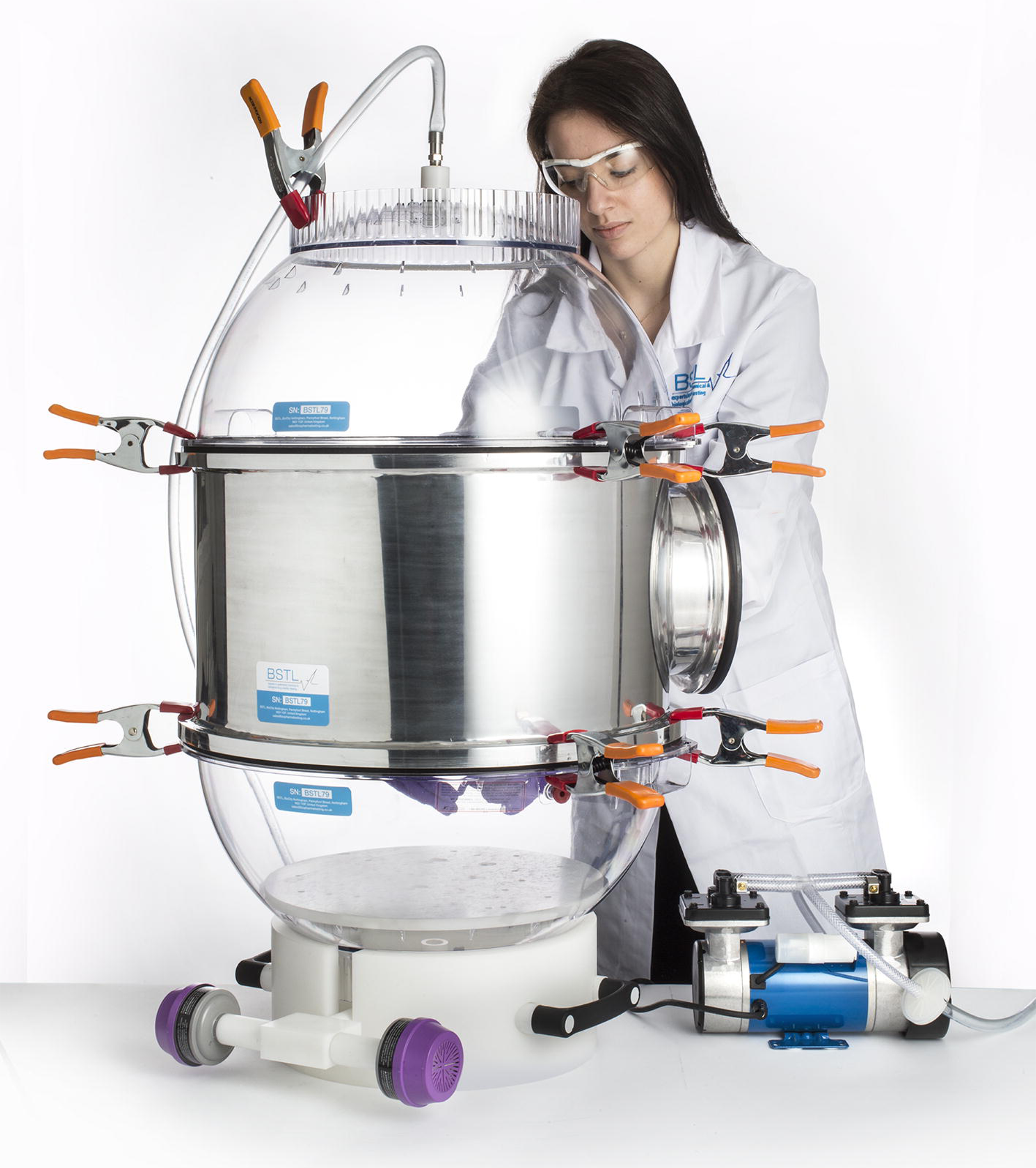
Biopharma Stability Testing Laboratory modified NIOSH chamber original copyright BSTL 2016, reprinted under a CC BY license, with permission from BSTL ltd.

## Results

Previously reported breakthrough tests confirmed no breakthrough of 2-POE using the air sampling approach described using Tenax TA sorbent and was not repeated in the present study [21]. Furthermore, recoveries for 2-POE of >99.9% from the Tenax TA air sampling tubes were demonstrated showing complete desorption of 2-POE. The air-sampling conditions during testing were safely within the upper limits for breakthrough for the Tenax sorbent media. System blank tubes analyzed after each test sample gave no detectable levels of 2-POE and hence demonstrated no carryover in the system. Using the analytical method described, an instrumental LOD of less than 0.3 ng for 2-POE and an experimental LLOQ of 0.41±0.01 ppbv were demonstrated. The lowest standard used for calibration was 0.6ng 2-POE due to the poorer recovery values at 0.3ng. The study investigated the containment performance of two physical barrier CSTDs (BD Phaseal™ and ChemoLock™) and two air-cleaning CSTDs (Tevadaptor™/ OnGuard™ and ChemoLock ™) using the 2016 Draft NIOSH universal CSTD test protocol [15]. The containment results data is shown below in table 2.

**Table 2.** CSTD containment performance data using 2.5% w/v 2-phenoxyethanol (2-POE) as challenge agent according to the 2016 Draft NIOSH universal CSTD test protocol for both tasks: task 1 (n = 4) and task 2 (n = 4).

| DEVICE TYPE | NIOSH TASK | MEAN CONCENTRATION OF 2-POE ± 95% CI (PPBV), N=4 | CALCULATED LLOQ (PPBV) |
| --- | --- | --- | --- |
| CHEMOLOCK™ BARRIER | 1 | <LLOQ (0.08 ± 0.02) | 0.41± 0.01 |
| CHEMOLOCK™ AIR-CLEANING | 1 | <LLOQ (0.10 ± 0.02) | 0.41± 0.01 |
| TEVADAPTOR™/ ONGUARD™ AIR-CLEANING | 1 | <LLOQ (0.03 ± 0.01) | 0.41± 0.01 |
| BD PHASEAL™ BARRIER | 1 | <LLOQ (0.05 ± 0.01) | 0.41± 0.01 |
| CHEMOLOCK™ BARRIER | 2 | <LLOQ (0.11 ± 0.05) | 0.41± 0.01 |
| CHEMOLOCK™ AIR-CLEANING | 2 | <LLOQ (0.19 ± 0.10) | 0.41± 0.01 |
| TEVADAPTOR™/ ONGUARD™ AIR-CLEANING | 2 | <LLOQ (0.04 ± 0.00) | 0.41± 0.01 |
| BD PHASEAL™ BARRIER | 2 | <LLOQ (0.05 ± 0.00) | 0.41± 0.01 |

CSTD: closed system transfer device; NIOSH: National Institute for Occupational Safety and Health; ppbv: parts per billion volume. Limit of detection (LOD) and lower limit of quantitation (LLOQ) were determined as 0.16 ± 0.01 ppbv and 0.41 ± 0.01 ppbv respectively which are significantly lower than previously published values [21]; CI: 95% confidence interval; Range is from the arithmetic mean minus the 95% CI (low) to the arithmetic mean plus the 95% CI (high).

The same number of replicates were used in this study (n = 4) as per the 2016 Draft NIOSH protocol (n = 4) and previous publication by Wilkinson *et al.* for consistency and comparison of test data [15,21]. All of the CSTDs evaluated gave below LLOQ releases of 2-POE and so performed on parity in terms of containment when 2.5% w/v 2-POE in water is used as challenge agent. This test included an equal number of physical barrier and air-cleaning CSTDs assessed using the 2016 Draft NIOSH universal CSTD test protocol in a head-to-head study. The 2016 Draft NIOSH universal protocol provides the ability to perform a direct comparison of containment performance between disparate CSTD technologies namely physical barrier and air-cleaning, the two main CSTD architectures in the market [15].

Chamber blank measurements (n = 2) were performed prior to each device test as described in the modified 2016 Draft NIOSH test protocol [15]. No statistically significant background contamination of 2-POE was present (data not shown) above nominal background levels for 2-POE which were on average 0.05±0.01 ppbv for 2-POE (n=85). A typical total ion gas chromatogram (TIC) separation of a chamber air sample containing a positive release of liquid 2-POE (2.5 % w/v in water) with detection by mass spectrometry is shown below in figure 4. The internal standard d8-toluene has a retention time of 9.79 minutes on column (blue trace A) and the analyte 2-POE has a retention time of 22.97 minutes on column (blue trace B) as shown by the extracted selected mass ions at M/Z 98 (d8-toluene) and M/Z 94 (2-phenoxyethanol).

**Figure 4.**
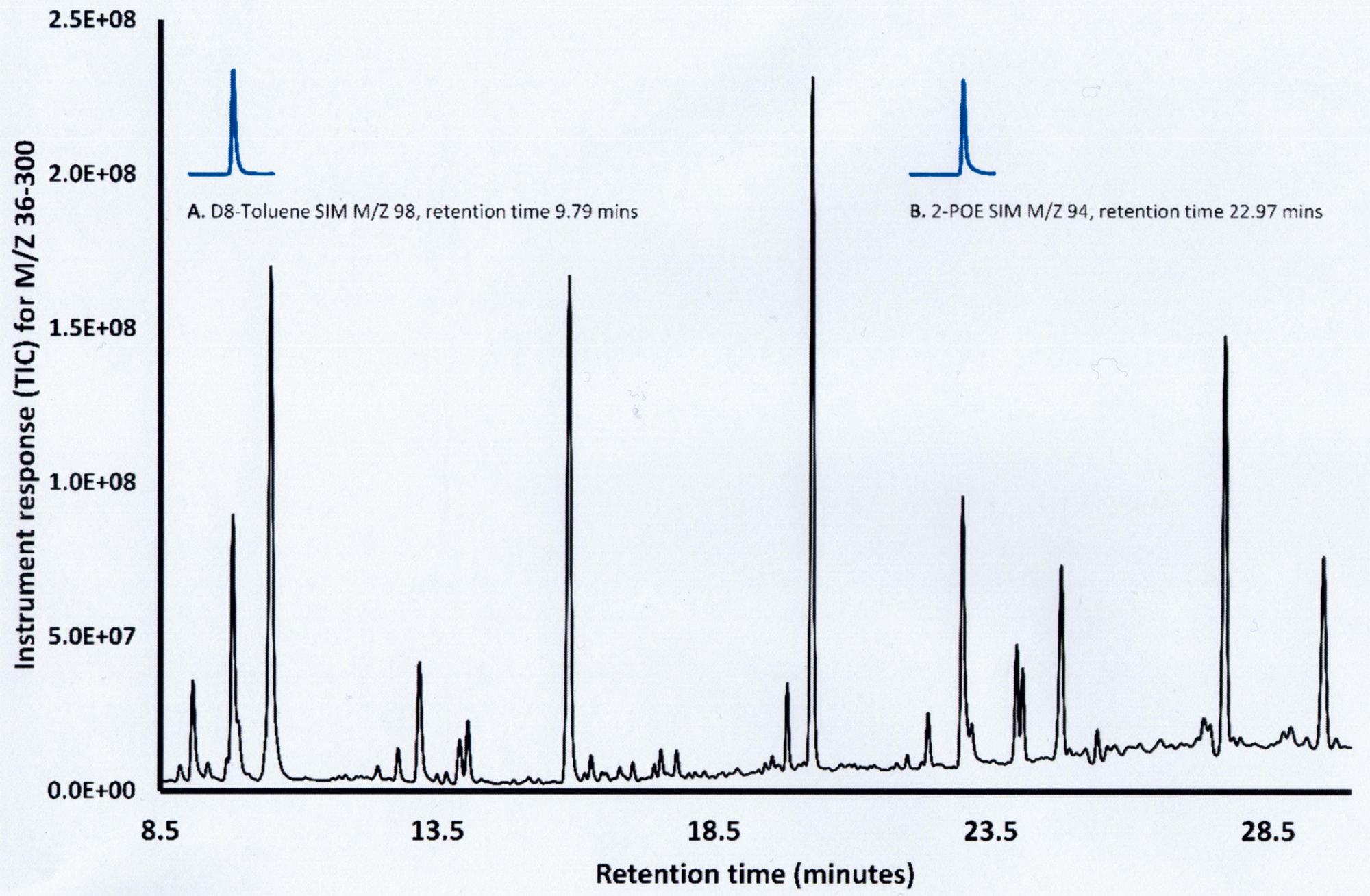
Typical total ion gas chromatogram (TIC) (black trace) of a positive liquid release of 2-phenoxyethanol in chamber air with mass spectrometry (extracted selected ion mases at M/Z 98 and 94 for d8-toluene and 2-phenoxyethanol, SIM traces shown in blue A and B respectively) detection.

A stacked plot was used in figure 4 to allow the SIM signal (blue) to be presented on the same ordinate axis (with an offset) as the total ion chromatogram (TIC) signal (black). The SIM plots (blue traces A and B) show the analyte and internal standard peaks only whereas the TIC chromatogram (black) shows all of the compounds collected on the Tenax tube during air sampling. It can be seen from the TIC chromatogram that there are a number of species present in the environmental NIOSH chamber air which are physically separated from the internal standard and analyte peaks by the measurement system.

In addition to the tests using the 2.5% w/v 2-phenoxyethanol challenge agent each CSTD was also evaluated for both task 1 and task 2 using 100% Milli-Q water as challenge agent without replication (n=1) as a negative control test. The data is shown above in table 3. The data shows values for 2-POE that are indistinguishable from the average blank value for 2-POE of 0.05±0.01 ppbv (n=85) demonstrating system suitability. No releases of 2-POE were obtained from either the test apparatus or CSTD devices and therefore releases of 2-POE from tests could have only originated from the 2.5% w/v 2-POE surrogate solution during testing.

**Table 3.** Data for the negative control test measurements obtained using 100% MilliQ water as the challenge agent for containment performance testing of the CSTD components tested under the 2016 Draft NIOSH universal test protocol Tasks 1 and 2.

| CSTD PRODUCT | NIOSH TASK | CONCENTRATION OF 2-POE (PPBV) FOR NEGATIVE CONTROL TESTS (N=1) |
| --- | --- | --- |
| **CHEMOLOCK™ BARRIER** | 1 | ≤LLOQ (0.04, n=1) |
| **CHEMOLOCK™ AIR-CLEANING** | 1 | ≤LLOQ (0.04, n=1) |
| **TEVADAPTOR™ / ONGUARD™**<br>**AIR-CLEANING** | 1 | ≤LLOQ (0.04, n=1) |
| **BD PHASEAL™ BARRIER** | 1 | ≤LLOQ (0.05, n=1) |
| **CHEMOLOCK™ BARRIER** | 2 | ≤LLOQ (0.06, n=1) |
| **CHEMOLOCK™ AIR-CLEANING** | 2 | ≤LLOQ (0.04, n=1) |
| **TEVADAPTOR™ / ONGUARD™**<br>**AIR-CLEANING** | 2 | ≤LLOQ (0.04, n=1) |
| **BD PHASEAL™ BARRIER** | 2 | ≤LLOQ (0.05, n=1) |

Positive control data for the deliberate liquid release of 5 and 10 microlitres of the 2.5% w/v 2-POE challenge agent during Task 1 yielded 2-POE releases of 3.13 ppbv and 6.59 ppbv (which is ∼8 and ∼16 x LLOQ respectively and therefore quantifiable). Positive control releases were significantly larger than the responses obtained during any of the CSTD testing which were all below LLOQ. The positive control results data demonstrated suitability of the system for detecting a relatively low liquid volume release of 2.5 %w/v 2-POE in the system estimated to be around 100 nanolitres of release by volume. It is also noteworthy that the system response for 2-POE appears to be proportional to the size of the liquid release of 2-POE which has previously been reported and is an important characteristic demonstrating the system suitability [21]. The measured signal scales proportionally with the physical volume of release of 2-POE.

The objective of the study was to determine if there were differences in containment performance between the two air-cleaning CSTD technologies and the two physical barrier CSTDs evaluated in the study. As can be seen from the containment data both air-cleaning platforms gave equivalent below LLOQ data in both NIOSH tasks 1 and 2, analogous to the physical barrier CSTDs. These data demonstrate that air-cleaning technologies have the capability to produce equivalent containment performance to physical barrier technologies when assessed using the 2016 Draft NIOSH universal test protocol and this represents a significant finding [15].

## Discussion

The present work as with previously published studies involving the use of 2.5% w/v 2-POE as surrogate have demonstrated the possibility to use a semi-volatile surrogate to obtain robust scientific data with extremely low LLOQ (sub-ppbv) which in the present study was determined as 0.41±0.01 ppbv [21]. Another facet of the 2016 Draft NIOSH universal test protocol is that release measurements for 2-POE are proportional to the amount of the liquid surrogate release as can be seen in figure 2, although releases cannot be differentiated by the type of leak *i.e* liquid, aerosol or vapour. This is an important development in terms of understanding containment performance of CSTDs and helps to better inform on health worker exposure.

The demonstrated ability of a containment performance test protocol to quantify very small liquid leaks on the nanolitre to microlitre volume scale is an important part of the system suitability as well as allowing comparison of releases between different device types *i.e* physical barrier and air-cleaning [15,21]. There are additional benefits to using 2-POE as the NIOSH surrogate as a consequence of the specificity that mass spectrometry affords when operated in SIM mode [21]. Mass spectrometry is the gold standard for detection and can unambiguously identify the surrogate based on selected ions only present in 2-POE giving extremely high specificity [21]. Other detection modalities such as spectroscopic techniques (of which infrared is one) offer much lower levels of specificity (as well as sensitivity) and as such the data obtained is of lower evidential quality [14].

All detection modalities must be capable of separating the analyte from any background chemical interference in the measurement domain. As can be seen from figure 4 (black chromatographic trace) the TIC which shows all masses of compounds separated on the GC column indicates that there are a plethora of potential interfering species. However, as the GC physically separates out/ removes interferants from reaching the MS detector coupled with the selective ion filters in the MS domain this leads to unambiguous determination of the surrogate 2POE [21]. GC-MS remains the gold standard for a raft of standard analytical methods published by the International Organization for Standardization [39], the Environmental Protection Agency, and OSHA for detecting vapors from volatile and semi-volatile chemicals [18,19].

The containment performance data obtained from this study using the Draft 2016 NIOSH universal CSTD test protocol showed that both physical barrier and air-cleaning CSTDs can demonstrate equivalent containment performance using 2.5% w/v 2-Phenoxyethanol in water as the challenge agent. This is an important finding as it shows that air-cleaning technology can offer parity in containment to physical barrier CSTD technology and is potentially as effective at protecting health workers against accidental exposure during hazardous drug preparation and administration. The containment data presented in this study quantifies contamination from the three main routes of release: (1) liquid, (2) aerosols and (3) vapour during the 2016 Draft NIOSH protocol related compounding (task 1) and administration (task 2) tasks.

Based on the extremely low LLOQ obtained of 0.41±0.01 ppbv our data demonstrates that both the physical barrier and air-cleaning CSTDs assessed in this study are able to contain the NIOSH surrogate 2-POE to below LLOQ under the test conditions of NIOSH task 1 and NIOSH task 2.

Furthermore, the two air-cleaning CSTDs showed equivalent containment performance down to the lowest levels of quantification for 2-phenoxyethanol suggesting that air-cleaning CSTDs can help to protect health workers from occupational exposure to the same extent as a physical barrier CSTD which is an important discovery.

There is currently no consensus on the choice of NIOSH surrogate from the list of nine proposed potential surrogates published in the 2016 Draft NIOSH universal protocol [15] and only Wilkinson *et al.* has previously published containment data on the use of two proposed NIOSH surrogates namely 2-POE and TEU [15,21–22].

In general, all semi-volatile materials will exhibit analogous behavior in terms of evaporation and generate similarly low airborne concentrations in the vapour based on the amount of material released and SVC although as Pengelly *et al.* and Wheeler have reported concentrations will be of order ∼1% of theoretical SVC [26,27]. As such there is no advantage of using 2-POE as NIOSH surrogate over any of the other NIOSH proposed surrogates although thus far the research findings of Pengelly *et al.* and Wilkinson *et al*. do not highlight any issues when using this proposed NIOSH surrogate from the 2016 Draft NIOSH protocol list [21,26]. For this work, surrogate selection foremost considered compatibility with common medical plastics which has been evaluated and demonstrated for 2-POE at 2.5% w/v (unpublished work). Finally for completeness it is important to mention two recent publications from Doepke *et al.* and Westbrook *et al.* which consider different highly volatile chemical surrogates to those proposed by the NIOSH in either the 2015 Draft NIOSH protocol or the 2016 Draft NIOSH universal test protocol documents [42,43]. A discussion of the latest research by Doepke *et al.* and Westbrook *et al.* is outside of the scope of the current study.

## Conclusions

In this study we report the containment performance of four commercial CSTD technologies in accordance with the 2016 Draft NIOSH universal CSTD test protocol and using 2.5 % w/v 2-phenoxyethanol as challenge agent following assessment in NIOSH Task 1 and Task 2 each performed in replicate (n=4) [15]. We selected two physical barrier and two air-cleaning CSTDs, which had not previously been assessed head-to-head using the 2016 Draft NIOSH universal test protocol [15]. The 2016 Draft NIOSH universal test protocol is referred to as the universal test protocol as it has the capability to assess and compare containment performance data from disparate CSTD technologies including both physical barrier and air-cleaning [15].

2-POE has previously proven to be an ideal challenge agent at 2.5 % w/v in water for testing of CSTDs [21]. Previously the LLOQ for 2-POE was established at 0.88 ppbv based on the average background blank chamber value which for the present study is 0.41±0.01 ppbv (LLOQ), almost half the value of the previous sensitivity level [21]. Thus, the testing presented here is significantly more sensitive (factor of two fold increase) in detecting potential releases of 2-POE than with the previous testing [21].

Both physical barrier CSTDs (ChemoLock™ and BD Phaseal™) the predicates, devices showed equivalent containment performance (at below LLOQ) to the two air-cleaning CSTDs evaluated (Tevadaptor™/ OnGuard™ and ChemoLock™ air-cleaning). As such the two physical barrier and two air-cleaning technologies can be regarded as being equivalent in terms of their potential to help reduce health worker exposure with all giving below LLOQ results.

However, it should be noted that extrapolation between disparate technologies that are manufactured by different companies is not possible. Each device technology needs to be evaluated according to the 2016 Draft NIOSH universal test protocol [15] due to manufacturing qualities and tolerances that are proprietary to each manufacturer for all safety devices and differences between manufacturers can be significant [14]. We can only report here that the CSTDs evaluated in this study demonstrated parity in containment performance with below LLOQ containment values. These findings underscore the importance of adopting standardised, semi-volatile surrogates as provided by the 2016 Draft NIOSH universal test protocol such as 2-POE and quantitative analytical methodologies such as TD-GC-MS to ensure reliable and comparable assessment of CSTD containment performance.

## Data Availability

All data produced in the present study are available upon reasonable request to the authors. All processed data produced in the present work are contained in the manuscript.

## Supporting Information

S1 Text. Supplementary materials

## Notes

### Competing Interest Statement

The authors have declared no competing interest.

### Funding

- ICU Medical (United States)

