## Supplemental Information for "Head-to-head containment performance testing of four commercially available closed-system transfer devices using the 2016 Draft NIOSH test protocol and 2.5% w/v 2-phenoxyethanol as challenge agent"

S1 Text. Supplementary Materials

Biopharma Stability Testing Laboratory (BSTL) Environmental Chamber

The BSTL chamber was manufactured according to the 2016 draft National Institute for Occupational Safety and Health (NIOSH) protocol [1] with the following modifications: The clear polycarbonate (PC) extension ring used in the NIOSH chamber was replaced with one constructed from aluminium that provided a more robust seal and was more resistant to ingress of organic materials. The changes made reduced the contamination burden of the chamber and simplified decontamination procedures following high releases of volatile organic challenge agents. The internal volume of the BSTL-modified NIOSH chamber is identical to the original NIOSH version at 125 litres. The aluminium provides an improved sealing capability due to the increased mass over the plastic extension piece and does not deform when pulled down using the hand clamps. The foam seal tape used by NIOSH was replaced by a continuous black neoprene gasket, dimensions 532 mm outer diameter x 506 mm internal diameter x 8 mm thick to British Standards BS2752 C40 (SJG international, UK). This change was made to improve the quality of the seal at each flange where the hemispherical domes mate to the middle extension piece. The change in materials does not have a detrimental effect on the performance of the test chamber and, if anything, improves the chemical resistance and the dimensional aspects of the seal. Internally the shelf that is supplied with the desiccator parts of the chambers (Belart, USA) was replaced by a Delrin shelf of the same dimensions but with an increased depth of 8 mm to improve structural rigidity during application of the vial adaptor to the drug vials. In the original design, the shelf flexed under the downward pressure necessary to perform the vial adaptor addition. All of the connecting fittings to the NIOSH chamber were manufactured with threaded parts and integral O-ring viton seals to ensure that a gas tight connection was maintained at all times. Figure 3. in the main body of the paper shows the BSTL-modified NIOSH chamber as used in this study.

Equipment

Stainless steel air sampling tubes (thermal desorption [TD tubes) were used with physical dimensions (89 mm (3.5”) long × 6.4 mm (¼”) outer diameter (OD) packed with 200 mg of Tenax TA sorbent, retained between two steel gauzes (Markes, UK). A TD tube conditioner (Markes TC-20, UK) was used to prepare tubes.

Sampling pumps (Gil-Air, UK) certified to BS EN ISO 13137 [2] and capable of maintaining a constant flow rate of 100 mL/min ± 5% were used. A calibrated flow meter with a measurement uncertainty of ± 2% was used to calibrate the flow rate of the air sampling pump prior to each use. A 1 cm length of silicone tubing (Fisher, UK) was used to connect the TD sampling tube to the pump.

The analytical equipment used in the analysis of the collected air samples is described below and comprises an automated Thermo Scientific Trace1300 gas chromatograph fitted with a TD100-xr thermal desorption autosampler (Markes, UK) interfaced to a Thermo Scientific ISQ 7000 AEI mass spectrometer, a VOCOL capillary column (60 m length × 0.25 mm diameter × 1 µm film thickness), and data was acquired and processed using the CDS software supplied by Thermo Scientific (Chromeleon version 7.3).

Chamber cleaning

Before first use, the chamber was completely dismantled, and each component thoroughly cleaned with 70% isopropyl alcohol (IPA)-water (Fisher, UK) and laboratory wipes containing no surfactants or additives followed by rinsing with Milli-Q water. This was repeated three times. After allowing the chamber to air dry, a ppbRAE 3000 photon-ionization detector (PID) (Shawcity, UK) was used to verify that no volatile residues were present on the surfaces of the chamber and or components.

Before each test the chamber was thoroughly cleaned according to a validated cleaning procedure ensuring no 2-phenoxyethanol (2-POE) is carried over between tests.

Chamber verification

Monthly chamber sealing verification checks were performed using a ppbRAE 3000 PID (Shawcity, UK) with parts per billion sensitivity to assure that the chamber had no leaks during the testing. This was performed using IPA as challenge agent. The chamber was assembled and sealed using four sprung hand clamps around the perimeter with the ppbRAE placed inside the chamber. An alcohol IPA swab was then opened outside the chamber and carefully used to trace along the points of contact between the hemispherical domes and the middle section as well as the gauntlet fixings and external top and bottom fittings to the chamber. The ppbRAE should not be able to detect any traces of IPA from inside the chamber if the sealing is effective.

Preparation of TD tubes prior to use

Pre-packed stainless steel air sampling tubes (TD tubes) were used with physical dimensions (89 mm [3.5”] long × 6.4 mm (¼”) outer diameter (OD) packed with 200 mg of Tenax TA sorbent, retained between two steel gauzes (Markes, UK). The tubes were fitted at both ends with metal storage end caps and PTFE ferrules and a unique identification number. Before use, the sample tubes were conditioned by heating in a flow of dry nitrogen in a tube conditioner (Markes TC-20, UK) at 300°C for 1 hour at a nominal flow rate of 70 mL/min. After conditioning, the tubes were cooled to room temperature and capped with the metal end caps. The sampling tubes were used and immediately prior to analysis spiked with an accurately known mass of *d_8_-*toluene according to a 1mL addition from a 1 parts per million (ppm) standard prepared in four nines di-nitrogen balance gas (BOC, UK).

Pre-test cleaning of the chamber

The gloves attached to the gauntlets were removed and disposed of prior to cleaning the chamber. To ensure that any traces of 2-POE were removed prior to each test, the chamber and gauntlets were thoroughly cleaned according to the following procedure: The internal shelf was removed and wiped with laboratory towel soaked with 70% IPA- Milli-Q water. Next, the shelf was washed with Milli-Q water and wiped dry with fresh laboratory towel. This was then repeated a second time and left to air dry. The two hemispherical domes of the chamber were placed uppermost and cleaned by wiping with a laboratory towel soaked in 70% IPA-Milli-Q water. A wash bottle was used to dispense copious amounts of Milli-Q water on to the surfaces of both hemispheres. Laboratory towels were then used to wipe the interior surfaces of the hemispheres working methodically from the top down. This was then repeated a second time and the hemispheres left to air dry. The gauntlets were wiped thoroughly with a laboratory towel soaked in 70% IPA-Milli-Q water but were not removed. The joining cuff rings and seal areas were cleaned carefully. The gauntlets were twice wiped using a laboratory towel soaked in Milli-Q water. The central aluminum section of the chamber was wiped using laboratory towel soaked in 70% IPA-Milli-Q water working from the top down. Milli-Q water was then used with a laboratory wipe to wash the central aluminum section working from the top down, which was repeated twice. All parts were left to air dry before replacing the internal shelf and re-assembly of the chamber including attachment of fresh nitrile gloves to the gauntlets. Finally, the air sampling pumps and trays were wiped over twice with a laboratory towel soaked in 70% IPA-Milli-Q water and then thoroughly wiped twice with a laboratory towel soaked in Milli-Q water and air dried before use. All test components to be tested in the chamber were handled with nitrile gloves to prevent contamination. Marker pens were avoided as these may contain 2-POE. The cleaning procedure was fully validated by BSTL using known amounts of 2-POE contamination inside the chamber. The cleaning procedure was able to remove all traces of 2-POE prior to collection of air samples in the post-clean blank samples. Air samples were collected in accordance with the HSE published methods for capture of volatile organic compounds [4].

Air sample collection

Duplicate air sampling pumps and sorbent tubes with loosened end caps were placed inside the chamber. If a test was being conducted, all test components were also placed inside the chamber. If a blank sample was collected, just the air sampling apparatus was introduced into the chamber. The chamber was then sealed using the four sprung hand clamps. The chamber was then flushed with a high flow rate (15 liters/min) of clean air by opening the inlet-air valve and activating an external B105 diaphragm pump (Charles Austen Pumps, UK) attached to the top outlet of the chamber. After 20 minutes, the pump was stopped and the inlet valve closed to seal the chamber. Next, the end caps were removed from the sorbent tubes and the latter was connected to the internal air sampling pumps. The pumps were activated and a 3 liter air sample collected over a 30 minutes period at 100 mL/min. The sorbent tubes were then capped and the chamber opened up. The sorbent tubes were placed in a Ziploc bag for storage prior to analysis. Air samples were collected in accordance with the HSE published methods for sampling of volatile organic compounds from the air [6].

Air sampling procedure

A sampling pump certified to BS EN ISO 13137 [2] and capable of maintaining a constant flow rate of 100 mL/min ± 5% was used (Gil-Air, UK). A calibrated flow meter with a measurement uncertainty of ± 2% was used to calibrate the flow rate of the air sampling pump prior to each use. A 1 cm length of silicone tubing was used to connect the sampling tube to the pump and was replaced between experiments.

For each test, air samples were collected using the TD tubes described, and a Gil-Air portable air sampling pump was set to deliver 100mL/min. A sampling time of 30 minutes was used giving a total air sampling volume of approximately 3 litres. Before and after sampling, the sampling tubes were capped with brass or aluminium end caps and locking nuts.

Disposable powder-free nitrile gloves were worn at all times when handling items to be placed in the chamber. These were changed frequently during the testing day and a fresh pair of gloves was used for every test.

Environmental background sample

An environmental background air sample was taken from the test laboratory environment outside the test chamber at the beginning of each test day. A single 3-litre air sample, collected as described for the testing, was used. In addition, and prior to collecting the environmental air sample, the presence of volatile organic compounds was checked using a ppbRAE 3000 PID (Shawcity, UK).

Chamber blank sample

Immediately prior to running a test sample, two chamber blank air samples were collected under conditions identical to the test samples. The blank samples were then used to calculate the experimental limit of detection (LOD).

Negative control sample

A single negative control test was always performed for each closed system transfer device (CSTD) tested for both NIOSH task 1 and task 2. This test was performed following the same procedure as for the CSTD sample tests but using drug vials containing 100% deionized (Milli-Q) water rather than the 2.5% w/v POE challenge agent.

Positive control sample

Positive samples were tested inside the chamber by carrying out the same NIOSH task 1 and task 2 test procedures but using an open system of needle and syringe as follows: A 21G x 1” needle was attached to one of the 60 mL Luer lock syringes. 45 mL of air was drawn up into the syringe before inserting the needle through the septum of drug vial 1. The vial was inverted, and the air was pushed into the vial, then liquid was allowed to enter the syringe during the equalization step ensuring that the tip of the needle stayed below the surface of the liquid in the vial at all times. The vial was returned to the upright position, and the needle and syringe were carefully removed. The 2.5% w/v 2-POE was transferred by inserting the needle through the vial septum of drug vial 2 and slowly injecting the liquid into the vial. At the end, an equal volume of air (45 mL) was removed prior to disconnecting the needle. The vial was swirled to simulate re-constitution. The needle was then re-inserted through the vial septum of vial 2, the vial invert and air injected into the vial, allowing 90 mL of liquid to enter the syringe before returning the vial to the upright position and removing the needle from the vial. For NIOSH task 1, the POE challenge agent (90 mL) was injected directly into the intravenous (IV) bag through the injection port of the bag. For NIOSH task 2, the needle was removed from the syringe and the syringe was directly attached to the Y-piece connector using the Luer lock fitting. The 90 mL of challenge agent was then added to the IV bag containing saline, with the tubing clamp open.

CSTD test procedure – NIOSH task 1 and 2

The CSTD test procedures were performed according to the 2016 draft NIOSH protocol [1] task 1 and 2. The manufacturer’s instructions for use (IFU) were followed for each CSTD for all manipulations. A total of five replicates were performed of each task.

Analytical conditions

The automated thermal desorption (ATD) conditions were as follows: valve temperature 225°C, desorb temperature 300°C, purge time 1 minute, desorption time 10 minutes, desorption flow rate 70 mL/min, trap temperature (low) −30°C, trap temperature (high) 270°C, outlet split flow 30 mL/min, column flow 1.3 mL/min, transfer line temperature 200°C. GC conditions were as follows: initial oven temperature 100°C (hold for 5 minutes), temperature ramp 1: 5°C/min (to 150°C; hold for 0 minutes), temperature ramp 2: 10°C/min (to 200°C; hold for 10 minutes). Under these conditions the retention times of internal standard *d_8_-*toluene and 2-POE were around 9.79 and 22.97 minutes, respectively. The MS conditions were as follows: multiplier voltage = 350 volts, total ion mass (low) = 25, total ion mass (high) = 200, selected ion masses for detection of *d_8_*-toluene = 95 – 105 m/z, selected ion masses for detection of 2-POE = 77, 94, and 138 m/z.

Instrument calibration

The calibration standards for analysis of 2-POE were prepared as follows: a stock solution of 2-POE was prepared by diluting approximately 20 milligrams of 2-POE (Sigma Alrich) to 100 mL with analytical grade methanol (Fisher, UK) to prepare a 200ng per microlitre primary stock solution. All volumes were accurately determined by mass using a 4-figure calibrated balance (Sartorius AC120S). Dilution of 0.5 mL of the stock solution to 0.5 mL with methanol provided standard 1. Dilution of 0.5 mL of the standard 1 to 0.5 mL with methanol provided standard 2. Dilution of 0.5 mL of standard 2 to 0.5 mL with methanol provided standard 3. Dilution of 0.5 mL of standard 3 to 0.5 mL with methanol provided standard 4. Dilution of 0.5 mL of standard 4 to 0.5 mL with methanol provided standard 5. Dilution of 0.5 mL of standard 5 to 0.5 mL with methanol provided standard 6. Dilution of 0.5 mL of standard 6 to 0.5 mL with methanol provided standard 7. Dilution of 0.5 mL of standard 7 to 2.5 mL with methanol provided standard 8. Dilution of 0.5 mL of standard 8 to 0.5 mL with methanol provided standard 9. 1 µl of each standard solution was then pipetted onto a separate Tenax TA tube using a calibrated micro-syringe. The transfer process was carried out in a flow of dry nitrogen (100 mL/min), which took 10 minutes to complete. The process described above generated a set of nine calibration tubes spiked with approximately 0.31, 0.63, 3.13, 6.25, 12.5, 25, 50, 100, and 200 ng of 2-POE and around 1500 ng of *d_8_-*toluene. The five standards were analyzed by ATD and GC-MS using the conditions described above and the selected ion peak areas of *d_8_-*toluene (*m/z* = 95 – 105) and 2-POE (*m/z* = 77, 94, and 138) were measured. The mean *d_8_-*toluene peak area obtained from the nine standards was calculated and used to normalize the 2-POE peak areas in each standard. The normalized peak areas were then used to construct a calibration graph, an example of which is shown in S1 Fig.

**S1 Fig.** Calibration graph for 2-phenoxyethanol (by ATD and GC-MS).

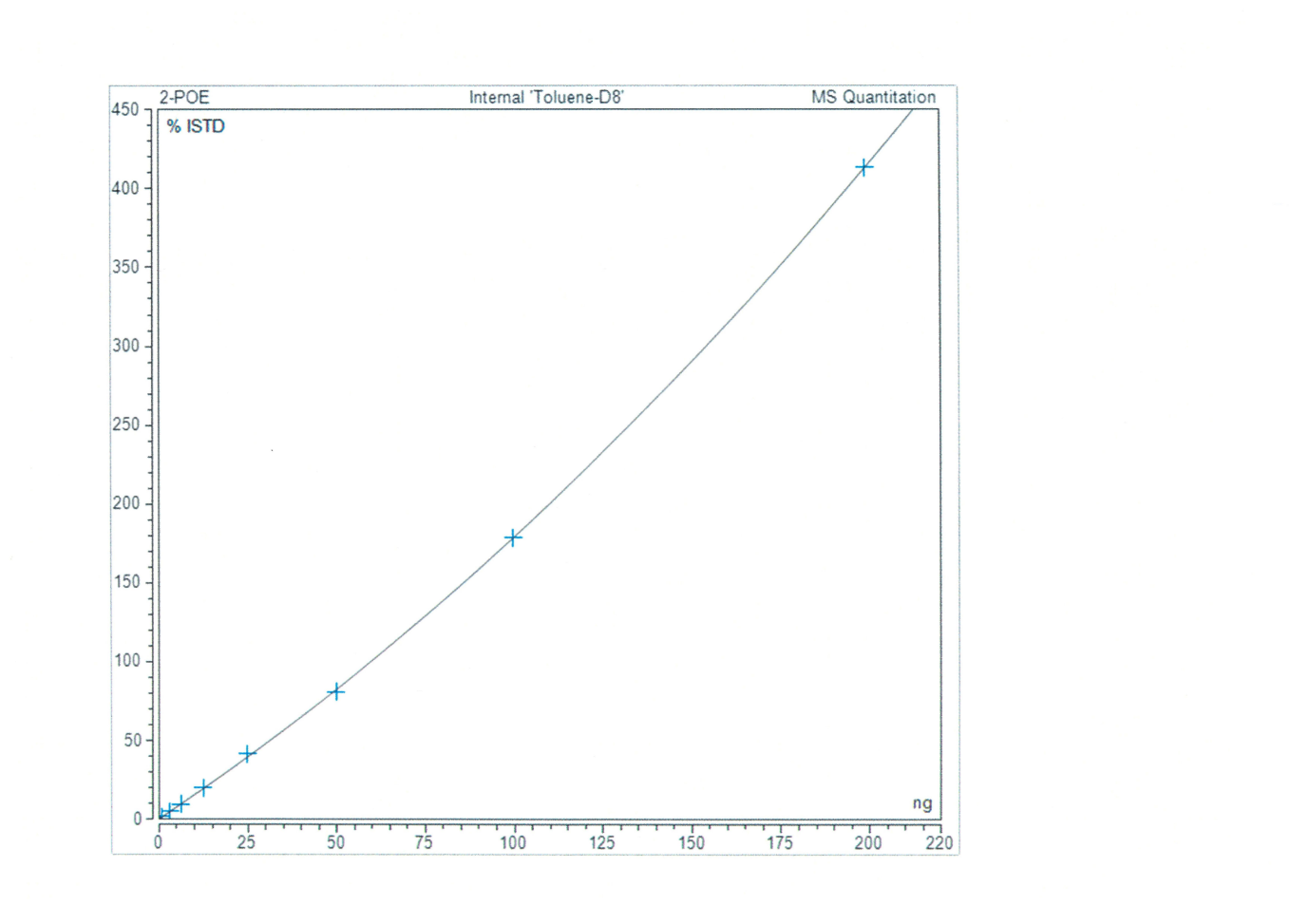

Sample Analysis

Sample tubes were analyzed using the conditions described above and the selected ion peak areas of *d_8_-*toluene (*m/z* = 95 – 105) and POE (*m/z* = 77, 94, and 138) were measured. The peak areas of the *d_8_-*toluene internal standard were then used to generate a normalized 2-POE peak area for each sample using equation 1.

**Equation 1:**	${POE}_{Norm}=P{OE}_{Sample}\times\frac{{D8}_{Mean}}{{D8}_{Sample}}$

where:

*POE_Norm_*	is the normalized peak area of 2-POE;

*POE_Sample_*	is the measured peak area for 2-POE;

*D8_Sample_*	is the measured peak area of *d_8_-*toluene and;

*D8_Mean_*	is the mean *d_8_-*toluene peak area in the five calibration standards.

The normalized peak areas were then converted to a mass of 2-POE, in ng, using the calibration graph prepared.

Determination of breakthrough volume and recovery

Breakthrough tests with sample volumes of 3, 6, and 12 liters were performed to verify that no breakthrough and >99.9% recovery from the Tenax TA occurred. Eight test tubes were pre-spiked with internal standard and 5 µl of standard 1, giving tube loading of ~180 ng of 2-POE and approximately 100 ng of *d_8_-*toluene. Two tubes were analyzed by TD-GC-MS immediately and then re-analyzed for traces of residual 2-POE in the second analysis. In between the first and second analysis, a clean blank tube was analyzed to demonstrate no carryover of 2-POE within the instrument and confirmed the trap desorption efficiency. The remaining six tubes were connected to a backup tube, loaded with internal standard but free of 2-POE. Dry air at 100 mL/min was pumped through the tubes for 30 (tubes 3 and 4), 60 (tubes 5 and 6), or 120 minutes (tubes 7 and 8), giving air sampling volumes of 3, 6, and 12-liters, respectively. At the end of the test, all twelve tubes were analyzed by TD-GC-MS and the results were used to confirm the system performance.

Evaporation test procedure

The following test procedure was developed to investigate the evaporation characteristics of any potential challenge agent and, in particular, to provide evidence of a proportional dependence between the volumes of challenge agent released into the chamber and the resulting airborne concentration of challenge agent that is measured. For the acceptable use of 2-POE as challenge agent, there must be a well-defined relationship between the amount of 2-POE released into the chamber and the concentration of 2-POE in the chamber air. The equation relating these two properties was found to be approximately linear over release values of between one and twenty-five microliters based on previous data. At higher release amounts, the relationship is non-linear reaching a maximum asymptotic value for 2-POE concentration. This may be due to the slower evaporative processes with larger drop sizes. For CSTD containment performance testing, a working range of one to twenty-five microliters is a useful dynamic range for assessment of liquid leaks. The chamber was first cleaned and used to collect a chamber blank (n = 2) as described, and the chamber was purged with clean air as described in the Methods section of the manuscript. Two air sampling tubes were uncapped, and the duplicate air sampling pumps were activated. A 5 µl aliquot of the challenge agent solution was dispensed onto a stainless steel metal surface inside the test chamber (using a Hamilton 10 microliter GC syringe). After 30 minutes, the air sampling pumps were switched off, the sorbent tubes were capped, and the chamber was opened. The procedure was then repeated using an aliquots of 10 µl of challenge agent solution. The sample tubes were analyzed to determine the airborne concentration of challenge agent in each test, and the results were plotted as concentration against volume of challenge agent solution dispensed. A linear relationship was observed between the volume of challenge agent dispensed inside the chamber and the airborne concentration of 2-POE inside the chamber (S2 Fig) consistent with low to moderate release amounts of 2-POE determined previously in the NIOSH chamber (1 to 25 µl). The relationship is non-linear at higher release values for 2-POE inside the chamber.

**S2 Fig.** Example plot for the evaporation of various volumes of 2.5% w/v aqueous 2-POE challenge agent solution in to the BSTL test chamber.

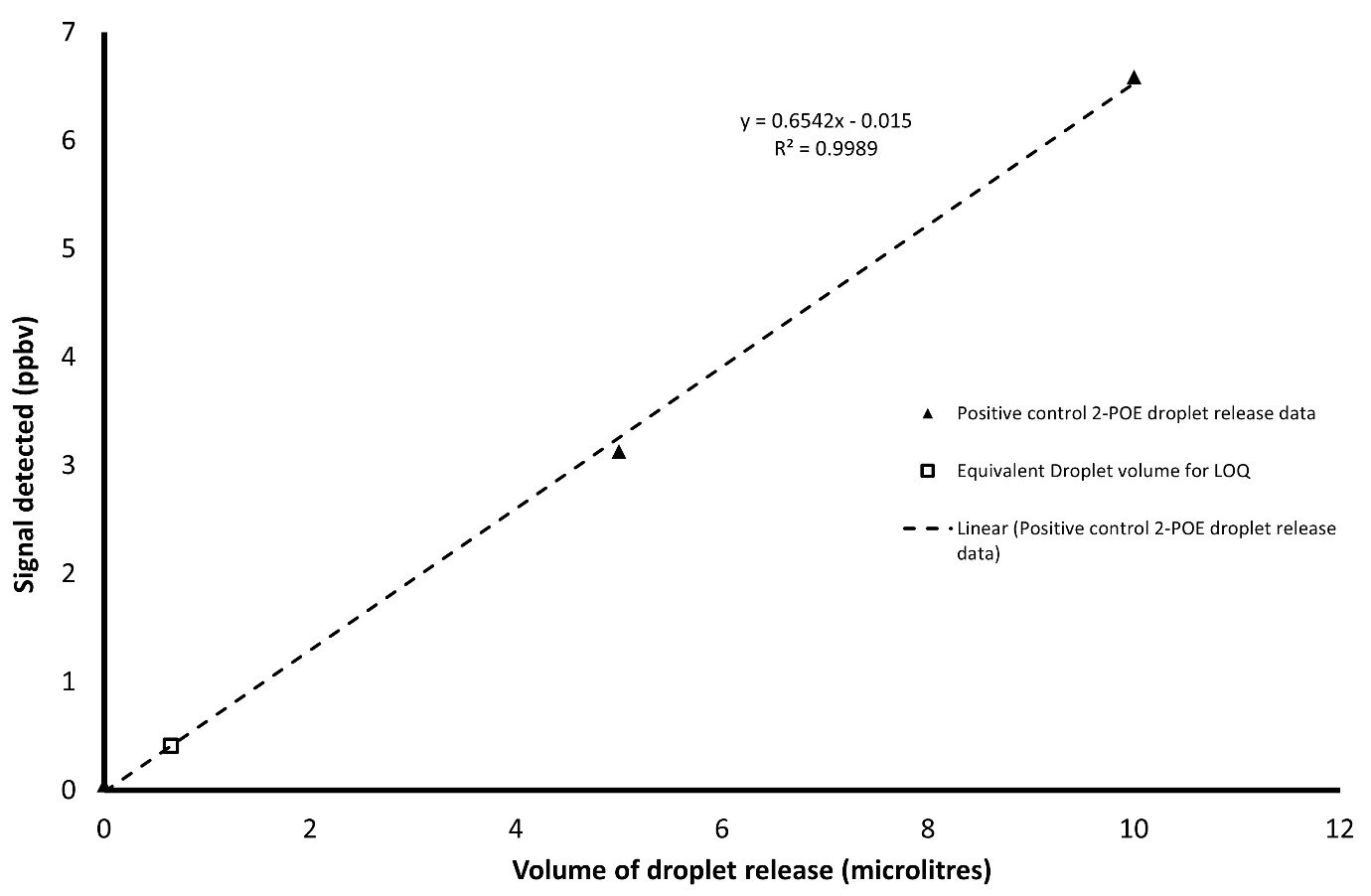
